# Open-source DeepSeek-R1 Outperforms Proprietary Non-Reasoning Large Language Models With and Without Retrieval-Augmented Generation

**DOI:** 10.1101/2025.09.12.25334809

**Authors:** Scott Song, Kenneth C. Peng, Elizabeth T. Wang, T.Y. Alvin Liu

## Abstract

**Objective:** To compare reasoning large language models (LLMs) vs. non-reasoning LLMs and open-source DeepSeek models vs. proprietary LLMs in answering ophthalmology board-style questions. To quantify the impact of retrieval-augmented generation (RAG).

**Design:** Cross-sectional evaluation of LLM performance before and after RAG integration.

**Subjects:** Seven LLMs: Gemini 1.5 Pro, Gemini 2.0 Flash, GPT-4 Turbo, GPT-4o, DeepSeek-V3, OpenAI-o1, and DeepSeek-R1.

**Methods:** A RAG-integrated LLM workflow was developed using the American Academy of Ophthalmology’s Basic and Clinical Science Course (*Section 12: Retina and Vitreous*) as an external knowledge source. The text was embedded into a Faiss vector database for retrieval. A curated set of 250 retina-related multiple-choice questions from OphthoQuestions was used for evaluation. Each model was tested under both pre-RAG (question-only) and post-RAG (question + retrieved context) conditions across 4 independent runs on the question set. Accuracy was calculated as the proportion of correct answers. Statistical analysis included paired t-tests, two-way ANOVA, and Tukey’s HSD test.

**Main Outcome Measures:** Accuracy (percentage of correct answers).

**Results:** RAG integration significantly improved accuracy across all models (p < 0.01). Two-way ANOVA confirmed significant effects of LLM choice (p < 0.001) and RAG status (p < 0.001) on model accuracy. Accuracy ranged from 56.8% (Gemini 1.5 Pro) to 87.5% (OpenAI-o1) pre-RAG, and improved post-RAG to 76.3% and 89.8%, respectively. Reasoning models (OpenAI-o1, DeepSeek-R1) significantly outperformed non-reasoning models. Open-source models achieved near parity with proprietary counterparts: DeepSeek-V3 with RAG (80.7%) performed comparably with GPT-4o with RAG (80.9%). DeepSeek-R1 with RAG slightly underperformed compared to OpenAI-o1 with RAG (86.0% vs 89.8%), but otherwise outperformed all other evaluated models (p < 0.001).

**Conclusion:** Our findings demonstrate that reasoning models significantly outperformed non-reasoning models, and RAG significantly enhanced accuracy across all models. Open-source models, trained at significantly lower cost, achieved near parity with proprietary systems. The performance of DeepSeek-V3 and DeepSeek-R1 highlighted the viability of cost-efficient, customizable, locally deployable LLMs for clinical applications. Future research should explore model fine-tuning, prompt engineering, and alternative retrieval methods to further improve LLM accuracy and reliability in medicine.

## Introduction

Large language models (LLMs) are artificial intelligence systems trained on vast amounts of text data, enabling them to generate human-like responses to prompts. In medicine, LLMs have been explored for clinical decision support, medical education, and patient communication, showing promise in assisting healthcare professionals.^1-4^ However, results have been mixed, with some studies highlighting knowledge gaps, misinformation, and inconsistent reliability, particularly in subspecialties requiring highly specific clinical knowledge.^4-5^

Within ophthalmology, LLMs have been evaluated for their responses to common patient inquiries and clinical case-centered inquiries.^6-9^ Previous studies have also assessed LLM performance on ophthalmology board-style questions, demonstrating variability across models and topic areas.^10-14^ A recent meta-analysis found that LLMs like GPT-4 outperformed earlier models but struggled with specific subspecialty areas, highlighting the knowledge constraints of LLM training data.^15^ Retrieval augmented generation (RAG) has emerged as a potential solution by incorporating external, authoritative knowledge sources to address LLM knowledge limitations.^16^ Studies implementing RAG frameworks in ophthalmology have demonstrated improved accuracy in answering subspecialty-specific questions.^17-19^ RAG-integrated models achieved higher accuracy than general-purpose LLMs by grounding responses in authoritative ophthalmic resources.^18^

While RAG frameworks address LLMs’ knowledge limitations by integrating external data, recently developed reasoning architectures represent an even more important advancement in LLM technology. In contrast to non-reasoning LLMs (e.g., GPT-4o, DeepSeek-V3), reasoning LLMs like OpenAI-o1 and DeepSeek-R1 employ reinforcement learning to simulate human-like, chain-of-thought reasoning chains.^20-21^ Unlike non-reasoning models that generate answers through rapid token prediction, reasoning models break complex queries into subproblems and synthesize intermediate conclusions before finalizing responses.^20-21^ This structured reasoning aligns closely with clinical decision-making processes where methodical analysis of evidence is critical, positioning reasoning LLMs as potential aids in diagnostics and treatment planning.

The recent release of powerful open-source LLMs like DeepSeek-V3 and DeepSeek-R1 has further expanded the potential of LLMs in medicine. These models combine state-of-the-art performance with significantly reduced computational costs, expanding access to high-performance LLMs for resource-constrained settings like hospitals.^21-22^ DeepSeek’s open-source architecture enabled tailored fine-tuning for medical domains, offering a scalable alternative to proprietary LLMs.^23-24^ However, a performance gap continues to persist between open-source models and proprietary systems like GPT-4, with past efforts narrowing, but not closing, this divide.^19^

This study aims to systematically evaluate reasoning versus non-reasoning models and open-source versus proprietary models in answering ophthalmology board-style questions, while evaluating the impact of RAG on LLM accuracy. Specifically, we included models, such as DeepSeek-V3 (non-reasoning, open-source), OpenAI-o1 (reasoning, proprietary), and DeepSeek-R1 (reasoning, open-source) to address the relative lack of published data on their medical capabilities in ophthalmology question-answering. Findings provide critical insights into the comparative advantages of reasoning models in clinical reasoning tasks, the viability of open-source models for clinical use, and RAG’s effectiveness in medical LLM applications.

## Methods

This study adheres to the principles of the Declaration of Helsinki and was reviewed by the Johns Hopkins IRB, which determined it to be exempt from further review.

This study evaluated the impact of RAG on the accuracy of LLMs in answering ophthalmology board-style questions. Five non-reasoning LLMs and two reasoning LLMs were included: Gemini 1.5 Pro (non-reasoning, proprietary), Gemini 2.0 Flash (non-reasoning, proprietary), GPT-4 Turbo (non-reasoning, proprietary), GPT-4o (non-reasoning, proprietary), DeepSeek-V3 (non-reasoning, open-source), OpenAI-o1 (reasoning, proprietary), and DeepSeek-R1 (reasoning, open-source). Each model was evaluated before and after RAG integration to compare performance.

### External Knowledge Base

The external knowledge base for RAG was *Section 12: Retina and Vitreous* of the 2023–2024 Basic and Clinical Science Course (BCSC) by the American Academy of Ophthalmology (AAO). This section provides comprehensive coverage of retinal physiology and pathology. As the gold standard for ophthalmology education, the BCSC serves as an ideal external knowledge source for enhancing LLM responses in board-style question-answering.

### Board-Style Question Curation

The question bank consisted of 250 text-only ophthalmology multiple-choice, board-style questions related to retina physiology and pathology, sourced from OphthoQuestions. Questions containing images or videos were excluded from the dataset.

### RAG-Integrated LLM Workflow

A RAG-integrated LLM workflow was developed in Python 3.11.8, leveraging the LangChain framework to orchestrate prompt generation, queries to OpenAI, Gemini, and DeepSeek LLMs, response processing, and RAG integration. To format the external knowledge base for RAG, text was split into overlapping 1,000-character chunks using LangChain’s *RecursiveCharacterTextSplitter*, embedded with Google’s *embedding-001* model, and indexed in a Faiss vector database for efficient retrieval.

### LLM Prompting and Evaluation

The latest versions of each LLM available at the time of data collection were used. Two custom prompt templates were designed for both pre-RAG and post-RAG conditions to standardize interactions with the LLMs, ensuring controlled comparisons and consistency in model prompting. Both templates included basic instructions requiring the same output format: a single-letter answer followed by an explanation. In the pre-RAG condition, the template combined these instructions with only the question stem. For the post-RAG condition, the template additionally incorporated context retrieved from the BCSC knowledge base, which was obtained by querying the local Faiss vector database (indexing BCSC text) with the question stem.

#### Pre-RAG Template

Instruction: Answer the following multiple-choice question by choosing the most correct answer between <<A>>, <<B>>, <<C>>, and <<D>>, and provide a 5-10 sentence explanation for why your answer is the most correct in comparison to the other answer choices.

Question: {question}

Answer:

#### Post-RAG Template

RAG Context: {context}

Instruction: Using both the provided context and your internal knowledge, answer the following multiple-choice question by choosing the most correct answer between <<A>>, <<B>>, <<C>>, and <<D>>, and provide a 5-10 sentence explanation for why your answer is the most correct in comparison to the other answer choices. In your explanation, make sure to explicitly cite any relevant information from the provided context that supports your selected answer.

Question: {question}

Answer:

To account for the inherent variance in LLM response generation, each LLM underwent 4 independent runs on the question bank for both pre-RAG and post-RAG conditions. Accuracy was calculated as the proportion of correctly answered questions, determined by evaluating the LLM responses against the question bank’s correct answers.

### Statistical Analysis

Statistical analyses were conducted using Python libraries, including scipy and statsmodels. Paired t-tests were used to assess the statistical significance of accuracy improvements between pre- and post-RAG conditions. A two-way analysis of variance (ANOVA) identified significant main effects of LLM choice (e.g., Gemini 2.0 Flash, GPT-4o, DeepSeek-R1), RAG status, and their interaction on accuracy. Post-hoc comparisons were performed using Tukey’s HSD test to explore pairwise differences. Statistical significance was defined as p < 0.05.

## Results

### Paired T-tests

In this study, we assessed the impact of RAG on the performance of seven LLMs in answering ophthalmology board-style questions. RAG integration led to a statistically significant accuracy improvement in all tested LLMs. Pre-RAG accuracy ranged from 56.8% for Gemini 1.5 Pro to 87.5% for OpenAI-o1, while post-RAG accuracy ranged from 76.3% for Gemini 1.5 Pro to 89.8% for OpenAI-o1 (Table 1). The accuracy improvements ranged from 19.5% for Gemini 1.5 Pro to 2.3% for OpenAI-o1. Paired t-tests demonstrated statistically significant improvements for all models (p < 0.01). The distribution of model accuracies is visualized in Figure 1. Post-RAG median accuracy values increased across all models, with the largest gains observed for Gemini 1.5 Pro and DeepSeek-V3.

**Table 1.** Comparison of model accuracy pre- and post-RAG integration. Accuracy scores reflect 4 test runs of each model on the same question set. Paired t-tests were used to evaluate whether the observed differences in accuracy with and without RAG were statistically significant (P < 0.05).

| Model Accuracy Comparison |  |  |  |
| --- | --- | --- | --- |
| LLM | Without RAG<sup>1</sup> | With RAG<sup>1</sup> | p-value<sup>2</sup> |
| Gemini 1.5 Pro | 56.8% (±0.73%) | 76.3% (±0.20%) | <0.001 |
| Gemini 2.0 Flash | 71.1% (±0.38%) | 78.4% (±0.57%) | <0.001 |
| GPT-4 Turbo | 69.3% (±1.15%) | 74.9% (±0.50%) | <0.001 |
| GPT-4o | 76.2% (±0.69%) | 80.9% (±0.76%) | <0.001 |
| DeepSeek-V3 | 70.2% (±0.23%) | 80.7% (±0.68%) | <0.001 |
| DeepSeek-R1 | 82.0% (±1.35%) | 86.0% (±0.65%) | 0.0016 |
| OpenAI-o1 | 87.5% (±0.38%) | 89.8% (±0.77%) | 0.0012 |

<sup>1</sup> Mean (± Standard Deviation)
<sup>2</sup> Paired t-test; P < 0.05 indicates statistical significance.

**Figure 1.**
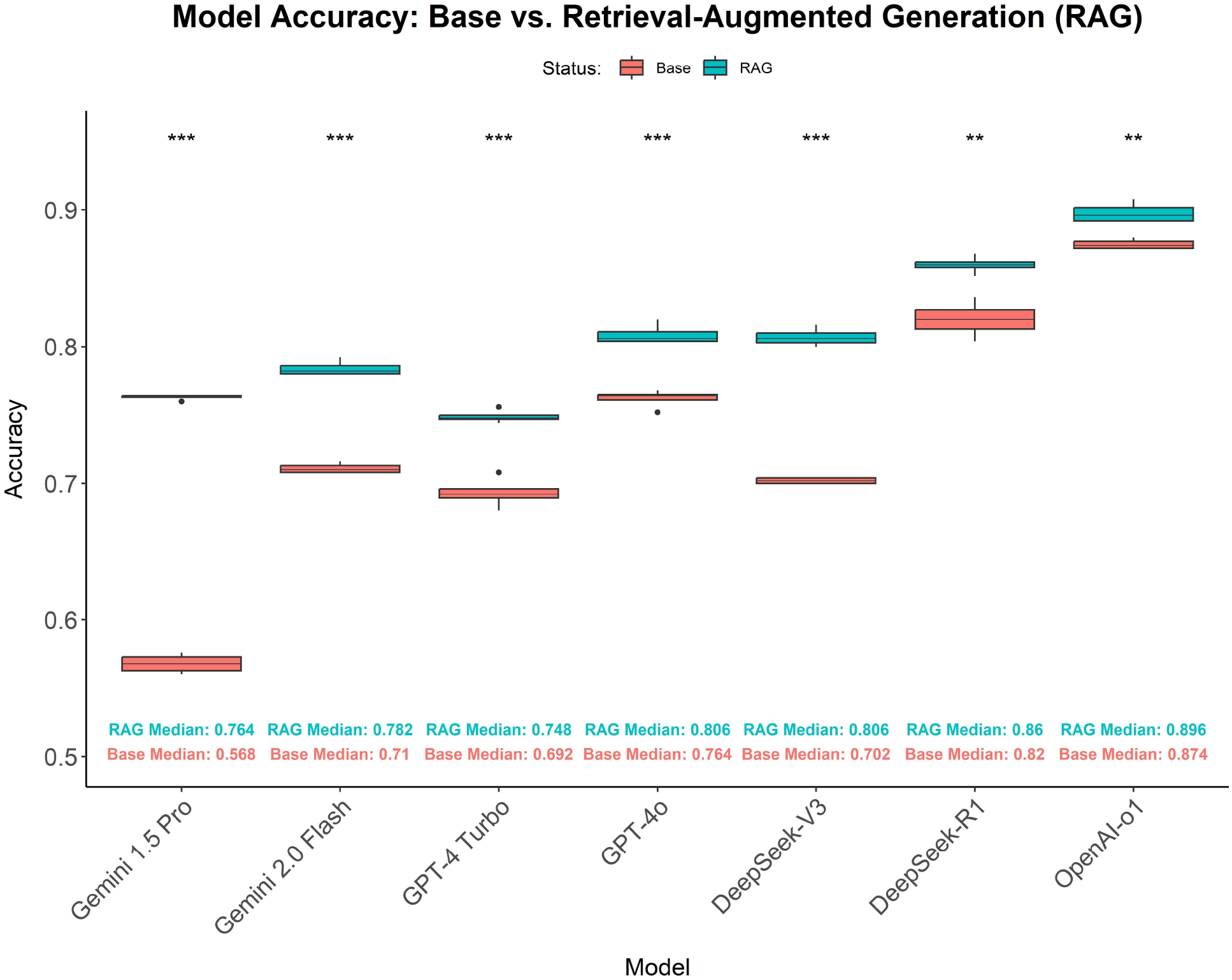
Box plot comparing model accuracies pre- and post-RAG integration. Data represents accuracy scores across 4 runs of a particular model on the same question set. Paired t-tests were used to evaluate whether the observed differences in accuracy with and without RAG were statistically significant (P < 0.05). *** = P < 0.001; ** = P < 0.01

### Two-way ANOVA

A two-way ANOVA identified significant main effects for LLM choice (p < 0.001) and RAG status (p < 0.001), as well as a significant interaction effect (p < 0.001) (Table 2). These findings indicate that both LLM type and RAG status significantly influence accuracy, with the magnitude of improvement varying across models.

**Table 2.**
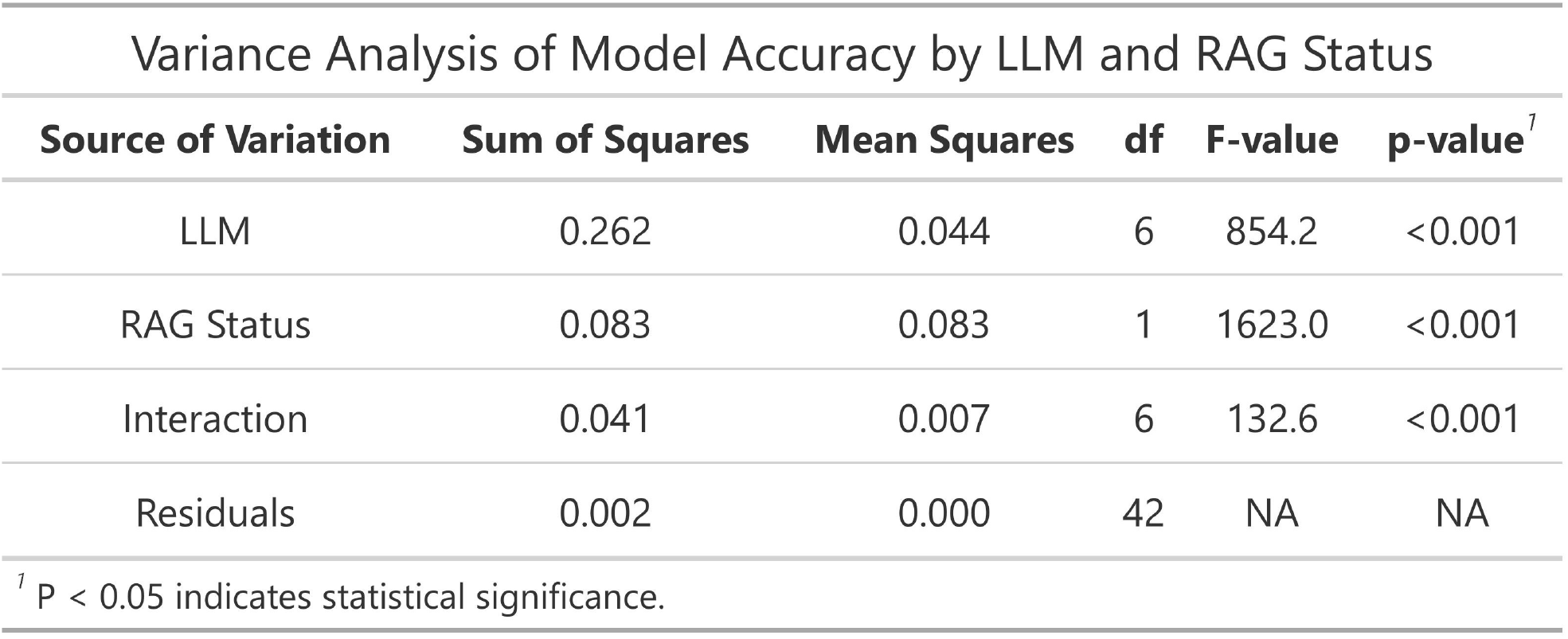
Results of a two-way analysis of variance examining the effects of LLM choice, RAG status, and their interaction on model accuracy. P < 0.05 indicates statistical significance.

### Tukey’s HSD Test

Tukey’s HSD test confirmed that RAG implementation significantly improved accuracy across all evaluated models (mean difference = 0.077, p < 0.001). Models with lower baseline accuracy, such as Gemini 1.5 Pro, benefited most from RAG integration, whereas higher-performing models like OpenAI-o1 and DeepSeek-R1 exhibited smaller but still statistically significant improvements. Specifically, Gemini 1.5 Pro improved from 56.8% to 76.3% (+19.5%; p < 0.001), Gemini 2.0 Flash improved from 71.1% to 78.4% (+7.3%; p < 0.001), GPT-4 Turbo improved from 69.3% to 74.9% (+5.6%; p < 0.001), GPT-4o improved from 76.2% to 80.9% (+4.7%; p <, DeepSeek-V3 improved from 70.2% to 80.7% (+10.5%; p < 0.001), and DeepSeek-R1 improved from 82.0% to 86.0% (+4.0%; p = 0.0016). OpenAI-o1, which had the highest baseline accuracy at 87.5%, showed the smallest relative gain (+2.3%, p = 0.0012) but maintained its position as the best-performing model with a post-RAG accuracy of 89.8%.

The mean accuracy differences between RAG-integrated models are shown in Figure 2. Collectively, RAG-integrated models significantly outperformed non-RAG models. DeepSeek-V3 with RAG outperformed Gemini 1.5 Pro, GPT-4 Turbo, and Gemini 2.0 Flash with RAG (p < 0.001). DeepSeek-V3 performed on par with GPT-4o with RAG (p = 1.0). Notably, DeepSeek-R1 significantly outperformed Gemini 1.5 Pro, Gemini 2.0 Flash, GPT-4 Turbo, GPT-4o, and DeepSeek-V3 in both pre-RAG and post-RAG configurations (p < 0.001). OpenAI-o1 significantly outperformed all evaluated models, including DeepSeek-R1, in both pre-RAG and post-RAG conditions (p < 0.001). However, only a small difference existed between DeepSeek-R1 with RAG and OpenAI-o1 with RAG (3.8%).

**Figure 2.**
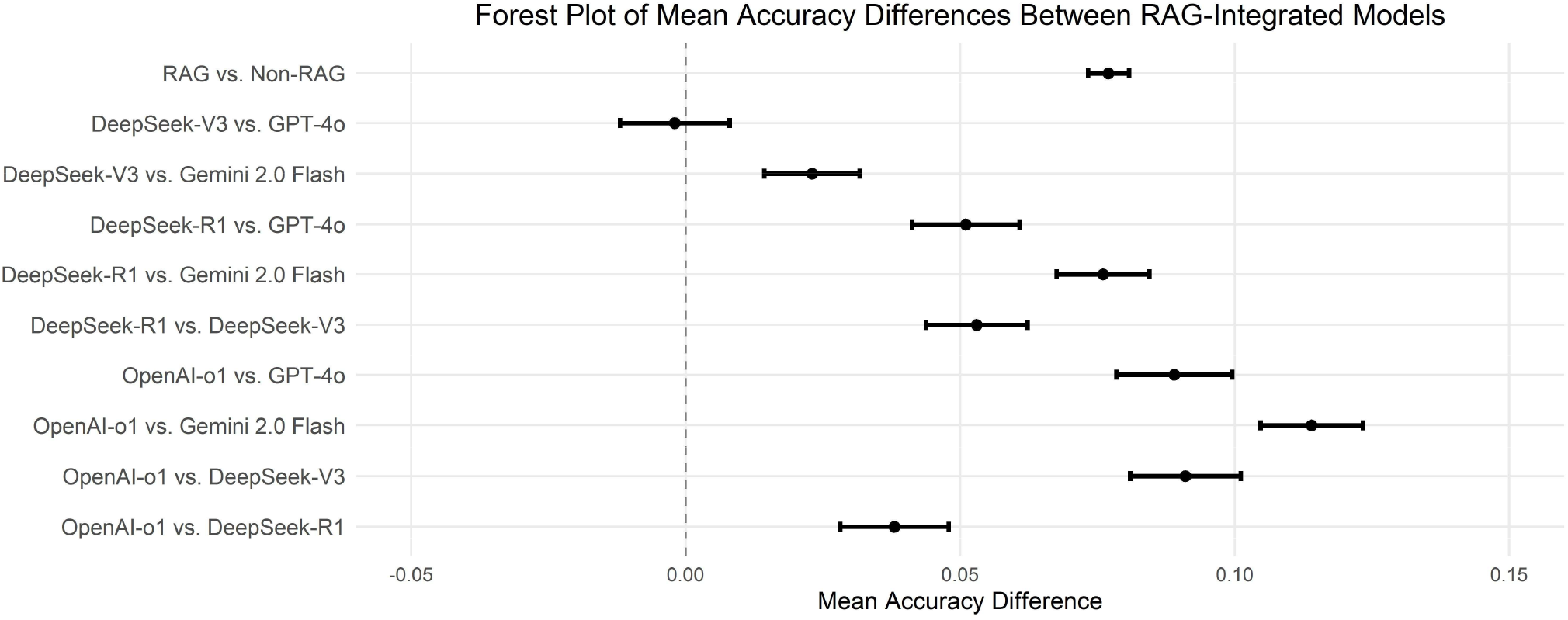
Forest plot of mean accuracy differences between RAG-integrated models. Error bars represent 95% confidence intervals for the difference in mean accuracy between RAG-integrated model pairs.

## Discussion

This study systematically compared proprietary versus emerging open-source models and reasoning versus non-reasoning models in answering ophthalmology board-style questions. Specifically, we included DeepSeek-V3 and DeepSeek-R1, two models that have gained significant attention in recent months due to their open-source nature and their significant lower cost of training. Our analysis revealed three key findings. First, reasoning models consistently outperformed non-reasoning models in both pre- and post-RAG conditions, highlighting the importance of chain-of-thought reasoning in clinical decision-making. Second, open-source models like DeepSeek-V3 and DeepSeek-R1 achieved near parity with proprietary models, despite being trained at a fraction of the cost, which is a promising finding, as this could accelerate democratization in access to high quality LLMs. In addition, our finding challenges the conventional thinking that proprietary models will always significantly outperform open-source models, especially if the performance gap continues to narrow. Third, despite baseline model accuracy ranging from 56.8% (Gemini 1.5 Pro) to 87.5% (OpenAI-o1), RAG significantly improved the performance of all evaluated models, reaffirming its value even as LLM capabilities advance. These findings highlight the transformative potential of reasoning architectures and open-source models in clinical applications while reinforcing RAG’s critical role in augmenting LLM accuracy.

One of the novelties in our current study is the inclusion of multiple reasoning LLMs. OpenAI-o1 and DeepSeek-R1 demonstrated significantly higher accuracy than all non-reasoning LLMs across both pre-RAG and post-RAG conditions (p < 0.001). Our finding aligns with OpenAI-o1’s state-of-the-art performance on the GPQA benchmark, a challenging dataset of multiple-choice questions that require advanced multi-step reasoning across scientific domains. Reasoning models like OpenAI-o1 outperformed non-reasoning models like GPT-4o by over 20% on the GPQA benchmark.^20^ Our study confirms that the superior performance of reasoning models seen in GPQA can likely be generalized to ophthalmology as well. We speculate that the capacity of reasoning models to decompose complex queries in GPQA evaluations mirrors clinical reasoning that is necessary in answering ophthalmology board-style questions. By employing methodical, chain-of-thought reasoning, reasoning models more closely emulate clinical decision-making processes. This architectural alignment with human reasoning may reduce hallucination risks and improve reliability in complex tasks, such as interpreting ambiguous symptoms or reconciling conflicting clinical recommendations. Furthermore, reasoning models’ capacity to generate intermediate logical steps enhances interpretability, allowing clinicians to audit the rationale behind responses, a critical feature for maintaining accountability in clinical workflows.

In recent months, one of the most exciting developments in LLMs is the rise in high-performing open-source models. Of these, DeepSeek-V3 and DeepSeek-R1 have gotten the most attention due to their exceptionally low cost in training. DeepSeek-V3, a non-reasoning LLM, achieved post-RAG performance (80.7%) comparable to GPT-4o (80.9%), while outperforming both Gemini 1.5 Pro and Gemini 2.0 Flash (p < 0.001). Furthermore, DeepSeek-R1, a specialized reasoning LLM, achieved an impressive post-RAG accuracy of 86.0%, surpassing all models evaluated in our study except for OpenAI-o1. Taken together, these results underscore the competitiveness of open-source models in specialized medical domains. This study is among the first in ophthalmology to demonstrate that open-source LLMs, leveraging both non-reasoning and reasoning architectures, can now achieve near-parity performance with proprietary LLMs. Beyond performance, open-source LLMs offer critical benefits for healthcare settings. First, they enable local hosting, eliminating the need to transmit sensitive patient data to third-party servers, thereby enhancing privacy compliance (e.g., HIPAA in the U.S.). Second, open-source models significantly reduce long-term costs compared to proprietary models, which often impose per-query fees. Third, they can be more easily adapted for clinical workflows by fine-tuning them on medicine-specific datasets, such as electronic health records, peer-reviewed literature, and institutional guidelines. Unlike proprietary models with rigid, opaque architectures and limited customization options, open-source models like DeepSeek can be readily fine-tuned to optimize their understanding of medical terminology, differential diagnoses, and real-world case scenarios. This flexibility enhances both accuracy and clinical relevance, ensuring their utility remains aligned with advancements in medical knowledge. By combining competitive performance with privacy, affordability, and adaptability, open-source models like DeepSeek-V3 and DeepSeek-R1 present a compelling alternative to proprietary models for medical institutions.

In terms of RAG, the observed accuracy improvements, ranging from 19.5% (Gemini 1.5 Pro) to 2.3% (OpenAI-o1), confirm the effectiveness of retrieval-based methods for enhancing model accuracy, supporting the growing body of evidence that LLMs benefit from grounding in curated medical knowledge.^16-19^ Beyond accuracy gains, RAG enhances clinical trustworthiness by enabling direct citations to external authoritative knowledge sources. For instance, while OpenAI-o1’s post-RAG accuracy improved modestly (87.5% to 89.8%), its responses became traceable to specific passages in Section 12 of the AAO’s BCSC and improved data provenance. This transparency is critical in clinical settings, where clinicians prioritize information from authoritative sources over ungrounded outputs.

This study has several limitations. First, the test set in our study only included 250 retina-specific multiple-choice questions, which may limit the generalizability of findings to other ophthalmic subspecialties. We decided to focus on questions from one subspecialty in order to streamline the curation process for our RAG external database. Second, our RAG external database was derived from the authoritative American Academy of Ophthalmology’s BCSC (*Section 12: Retina and Vitreous*). However, for future directions, we could include additional sources for external knowledge, such as peer-reviewed literature and clinical guidelines. Finally, the smaller accuracy gains observed in OpenAI-o1 and DeepSeek-R1 suggest a potential ceiling effect in higher-performing models, where additional knowledge retrieval yields diminishing returns. Future research should explore alternative retrieval strategies, embedding models, vector databases, and prompt engineering techniques to comprehensively evaluate RAG’s effectiveness in optimizing LLMs with high baseline performance. Additionally, studies should investigate whether fine-tuning RAG-integrated DeepSeek models can further enhance performance, particularly in complex medical reasoning and clinical decision-making. These research avenues may offer valuable insights into refining LLM applications in ophthalmology and other medical fields.

## Data Availability

All data produced in the present study are available upon reasonable request to the authors

**List of Abbreviations**
LLM: (Large language model)
RAG: (Retrieval-augmented generation)
BCSC: (American Academy of Ophthalmology’s Basic and Clinical Science Course)
ANOVA: (Analysis of Variance)
HSD: (Tukey’s honestly significant difference)

## Notes

**Conflict of Interest/Disclosures:** No conflicting relationship exists for any author.

### Competing Interest Statement

The authors have declared no competing interest.

### Funding Statement

This research was supported by Gills AI Center at the Wilmer Eye Institute and the Dean's Summer Research Fund from the Johns Hopkins University School of Medicine. Author TYAL was supported by the Research to Prevent Blindness Career Development Award. The sponsor or funding organizations had no role in the design or conduct of this research.

## References

1. Clusmann J, Kolbinger FR, Muti HS, et al. The future landscape of large language models in medicine. Commun Med (Lond). 2023;3(1):1–8. doi:10.1038/s43856-023-00370-1

2. Kung TH, Cheatham M, Medenilla A, et al. Performance of ChatGPT on USMLE: Potential for AI-assisted medical education using large language models. PLOS Digit Health. 2023;2(2):e0000198 . doi:10.1371/journal.pdig.0000198

3. Singhal K, Tu T, Gottweis J, et al. Toward expert-level medical question answering with large language models. Nat Med. Published online January 8, 2025:1–8. doi:10.1038/s41591-024-03423-7

4. Thirunavukarasu AJ, Ting DSJ, Elangovan K, Gutierrez L, Tan TF, Ting DSW. Large language models in medicine. Nat Med. 2023;29(8):1930–1940. doi:10.1038/s41591-023-02448-8

5. Singhal K, Azizi S, Tu T, et al. Large language models encode clinical knowledge. Nature. 2023;620(7972):172–180. doi:10.1038/s41586-023-06291-2

6. Lim ZW, Pushpanathan K, Yew SME, et al. Benchmarking large language models’ performances for myopia care: a comparative analysis of ChatGPT-3.5, ChatGPT-4.0, and Google Bard. EBioMedicine. 2023;95:104770. doi:10.1016/j.ebiom.2023.104770

7. Cheong KX, Zhang C, Tan TE, et al. Comparing generative and retrieval-based chatbots in answering patient questions regarding age-related macular degeneration and diabetic retinopathy. Br J Ophthalmol. 2024;108(10):1443–1449. doi:10.1136/bjo-2023-324533

8. Pushpanathan K, Zou M, Srinivasan S, et al. Can OpenAI’s New O1 Model Outperform Its Predecessors in Common Eye Care Queries? Ophthalmol Sci. 2025;0(0). doi:10.1016/j.xops.2025.100745

9. Huang AS, Hirabayashi K, Barna L, Parikh D, Pasquale LR. Assessment of a Large Language Model’s Responses to Questions and Cases About Glaucoma and Retina Management. JAMA Ophthalmol. 2024;142(4):371–375.

10. Antaki F, Touma S, Milad D, El-Khoury J, Duval R. Evaluating the Performance of ChatGPT in Ophthalmology: An Analysis of Its Successes and Shortcomings. Ophthalmol Sci. 2023;3(4):100324. doi:10.1016/j.xops.2023.100324

11. Thirunavukarasu AJ, Mahmood S, Malem A, et al. Large language models approach expert-level clinical knowledge and reasoning in ophthalmology: A head-to-head cross-sectional study. PLOS Digit Health. 2024;3(4):e0000341 . doi:10.1371/journal.pdig.0000341

12. Antaki F, Milad D, Chia MA, et al. Capabilities of GPT-4 in ophthalmology: an analysis of model entropy and progress towards human-level medical question answering. Br J Ophthalmol. 2024;108(10):1371–1378. doi:10.1136/bjo-2023-324438

13. Botross M, Mohammadi SO, Montgomery K, Crawford C. Performance of Google’s Artificial Intelligence Chatbot “Bard” (Now “Gemini”) on Ophthalmology Board Exam Practice Questions. Cureus. 2024;16(3):e57348 . doi:10.7759/cureus.57348

14. Gill GS, Tsai J, Moxam J, Sanghvi HA, Gupta S. Comparison of Gemini Advanced and ChatGPT 4.0’s Performances on the Ophthalmology Resident Ophthalmic Knowledge Assessment Program (OKAP) Examination Review Question Banks. Cureus. 2024;16(9):e69612 . doi:10.7759/cureus.69612

15. Wu JH, Nishida T, Liu TYA. Accuracy of large language models in answering ophthalmology board-style questions: A meta-analysis. Asia Pac J Ophthalmol (Phila). 2024;13(5):100106. doi:10.1016/j.apjo.2024.100106

16. Gao Y, Xiong Y, Gao X, et al. Retrieval-Augmented Generation for Large Language Models: A Survey. arXiv. Published online March 27, 2024. doi:10.48550/arXiv.2312.10997

17. Luo MJ, Pang J, Bi S, et al. Development and Evaluation of a Retrieval-Augmented Large Language Model Framework for Ophthalmology. JAMA Ophthalmol. 2024;142(9):798–805. doi:10.1001/jamaophthalmol.2024.2513

18. Singer MB, Fu JJ, Chow J, Teng CC. Development and Evaluation of Aeyeconsult: A Novel Ophthalmology Chatbot Leveraging Verified Textbook Knowledge and GPT-4. J Surg Educ. 2024;81(3):438–443. doi:10.1016/j.jsurg.2023.11.019

19. Nguyen Q, Nguyen DA, Dang K, et al. Advancing Question-Answering in Ophthalmology with Retrieval-Augmented Generation (RAG): Benchmarking Open-source and Proprietary Large Language Models. medRxiv. Published online November 19, 2024:2024.11.18.24317510. doi:10.1101/2024.11.18.24317510

20. Learning to reason with LLMs. Accessed March 16, 2025. https://openai.com/index/learning-to-reason-with-llms/

21. DeepSeek-R1 Release | DeepSeek API Docs. Accessed February 24, 2025. https://api-docs.deepseek.com/news/news250120

22. Introducing DeepSeek-V3 | DeepSeek API Docs. Accessed February 24, 2025. https://api-docs.deepseek.com/news/news1226

23. DeepSeek-AI, Liu A, Feng B, et al. DeepSeek-V3 Technical Report. Published online February 18, 2025. doi:10.48550/arXiv.2412.19437

24. DeepSeek-AI, Guo D, Yang D, et al. DeepSeek-R1: Incentivizing Reasoning Capability in LLMs via Reinforcement Learning. Published online January 22, 2025. doi:10.48550/arXiv.2501.12948

